# Markers Of Coagulation And Hemostatic Activation Identify COVID-19 Patients At High Risk For Thrombotic Events, ICU Admission and Intubation

**DOI:** 10.1101/2020.10.04.20206540

**Authors:** Darwish Alabyad, Srikant Rangaraju, Michael Liu, Rajeel Imran, Christine L. Kempton, Milad Sharifpour, Sara C. Auld, Manila Gaddh, Roman Sniecinski, Cheryl L. Maier, Jeannette Guarner, Alexander Duncan, Fadi Nahab

## Abstract

**Background:** Coronavirus disease 2019 (COVID-19) has been associated with a coagulopathy giving rise to venous and arterial thrombotic events. The objective of our study was to determine whether markers of coagulation and hemostatic activation (MOCHA) on admission could identify COVID-19 patients at risk for thrombotic events and other complications.

**Methods:** COVID-19 patients admitted to a tertiary academic healthcare system from April 3, 2020 to July 31, 2020 underwent standardized admission testing of MOCHA profile parameters (plasma d-dimer, prothrombin fragment 1.2, thrombin-antithrombin complex, and fibrin monomer) with abnormal MOCHA defined as ≥ 2 markers above the reference. Prespecified thrombotic endpoints included deep vein thrombosis, pulmonary embolism, myocardial infarction, ischemic stroke, and access line thrombosis; other complications included ICU admission, intubation and mortality. We excluded patients on anticoagulation therapy prior to admission and those who were pregnant.

**Results:** Of 276 patients (mean age 59 ± 6.4 years, 47% female, 62% African American race) who met study criteria, 45 (16%) had a thrombotic event. Each coagulation marker on admission was independently associated with a vascular endpoint (p<0.05). Admission MOCHA with ≥ 2 abnormalities (n=203, 74%) was associated with in-hospital vascular endpoints (OR 3.3, 95% CI 1.2-8.8), as were admission D-dimer ≥ 2000 ng/mL (OR 3.1, 95% CI 1.5-6.6), and admission D-dimer ≥ 3000 ng/mL (OR 3.6, 95% CI 1.6-7.9). However, only admission MOCHA with ≥ 2 abnormalities was associated with ICU admission (OR 3.0, 95% CI 1.7-5.2) and intubation (OR 3.2, 95% CI 1.6-6.4), while admission D-dimer ≥2000 ng/mL and admission D-dimer ≥ 3000 ng/mL were not associated. MOCHA and D-dimer cutoffs were not associated with mortality. Admission MOCHA with <2 abnormalities (26% of the cohort) had a sensitivity of 88% and negative predictive value of 93% for a vascular endpoint.

**Conclusions:** Admission MOCHA with ≥ 2 abnormalities identified COVID-19 patients at increased risk of ICU admission and intubation during hospitalization more effectively than isolated admission D-dimer measurement. Admission MOCHA with <2 abnormalities identified a subgroup of patients at low risk for vascular events. Our results suggest that an admission MOCHA profile can be useful to risk-stratify COVID-19 patients.

## INTRODUCTION

Coronavirus disease 2019 (COVID-19) has been widely associated with the development of a systemic coagulopathy that is likely multifactorial in etiology.^1^ COVID-19-associated coagulopathy exhibits the classic components of Virchow’s triad, and there are ongoing efforts to better characterize the underlying pathophysiology of the hypercoagulable state in the setting of COVID-19 infection.^2^ Elevated levels of fibrin degradation products, particularly D-dimer, have been used to predict venous thromboembolic events (VTE) and hypercoagulable states in various disease states.^3,4^ Plasma D-dimer in COVID-19 patients was found to be a useful predictor of VTE^5,6^ and thresholds of 2000 ng/mL or 3000 ng/mL have been suggested as indicators of disease severity.^7,8^

A biomarker panel called the markers of coagulation and hemostatic activation (MOCHA), which includes plasma D-dimer, prothrombin fragment 1.2, thrombin-antithrombin complex and fibrin monomer levels, has been shown to predict subsequent diagnosis of new malignancy, VTE, and other hypercoagulable states in patients with cryptogenic stroke.^9,10^ A previous study has also demonstrated that a MOCHA profile with < 2 marker abnormalities can effectively rule out hypercoagulable states in patients with embolic stroke of undetermined source.^11^

The objective of this study was to determine whether the admission MOCHA profile in hospitalized COVID-19 patients could aid in risk stratification by identifying those patients at low versus high risk of vascular thrombotic events, ICU admission, intubation and clinical outcome more effectively than D-dimer alone.

## METHODS

### Study Design and Setting

Patients were prospectively identified from an admission census list that identified all COVID-19 patients admitted to four hospitals in Emory Healthcare, an urban, academic, tertiary healthcare system in Atlanta, Georgia who had a MOCHA profile ordered on admission as part of a standardized COVID-19 orderset from April 3, 2020 through July 31, 2020. Expert consensus of a multidisciplinary working group recommended obtaining a MOCHA profile on all patients admitted with a diagnosis of COVID-19. All patients were 18 years of age or older with diagnosis of COVID-19 confirmed by PCR and had a MOCHA profile drawn within 72 hours of hospitalization. For this analysis we excluded patients on outpatient anticoagulation therapy prior to hospitalization due to the effect of anticoagulation on the MOCHA profile^10^ and pregnant patients due to the lack of validated reference ranges. Electronic medical records were retrospectively reviewed from admission through discharge or until the censor date of September 14, 2020 to identify venous and arterial thrombotic events. This study was approved by the Emory University Institutional Review Board.

Patient demographics including age, sex, race, body mass index (BMI), and a history of comorbidities including smoking, diabetes, hypertension, asthma, chronic obstructive pulmonary disorder (COPD), human immunodeficiency virus (HIV) infection, end stage renal disease, atrial fibrillation, prior VTE, coronary artery disease (CAD), stroke, and known active cancer were collected. Total length of stay, length of intensive care unit (ICU) admission, length of intubation, and final disposition were additionally recorded. Prespecified venous and arterial endpoints monitored during hospitalization included deep vein thromboses (DVT), pulmonary embolus (PE), myocardial infarction (MI), ischemic stroke, and dialysis or central line clots. DVT was confirmed by duplex ultrasound and PE by CT, CT angiography or ventilation/perfusion scans. MI was diagnosed as a troponin elevation and confirmation by a board-certified cardiologist. Ischemic stroke was diagnosed by CT or MRI and confirmation by a board-certified neurologist. Central access and renal replacement therapy circuit thrombosis were determined based on chart documentation.

### Laboratory testing

Admission MOCHA profiles were obtained within 72 hours of hospitalization and included plasma levels of D-dimer (reference value <574 ng/mL), prothrombin fragment 1.2 (reference range 65-288 pmol/L), thrombin-antithrombin complex (reference range 1.0-5.5 µg/L), and fibrin monomer (reference value <7 µg/mL). All assays were performed in the hospital clinical laboratories using 3.2% citrated plasma specimens. D-dimer levels were measured with high-sensitivity latex dimer assay (Instrumentation Laboratories, Bedford, MA). Both prothrombin fragment 1.2 and thrombin-antithrombin complexes were measured with the Enzygnost ELISA kit (Siemens Healthcare, Tarrytown, NY). Soluble fibrin monomer levels were measured by latex immunoassay (Stago, Parsippany, NJ).

### Statistical Analysis

Descriptive statistics were used to summarize the data with results reported as percentages for categorical variables. Means with standard deviations or medians with interquartile ranges were reported for normally distributed and skewed variables, respectively. Comparisons between means or medians of continuous variables were assessed with independent T-tests (two-tailed) or by the Mann-Whitney U non-parametric test, respectively. Categorical variables were compared with Pearson’s chi-square and Fisher Exact tests. Significance for all descriptive analyses was set at p<0.05. Univariable analyses were conducted using binary logistic regression for binary outcome variables (vascular endpoint, VTE, ICU admission, intubation and mortality).

Statistically significant predictors of these outcomes (p<0.10) were then considered in multivariable binary logistic regression analyses to identify independent outcome predictors (adjusted p<0.05). The association with thrombotic events within 14 days of admission was additionally examined.

The frequency of D-dimer and MOCHA profile abnormalities were recorded and subsequently analyzed in specific patient subsets, including those with ICU admission and intubation. The four parameters of the MOCHA profile were independently assessed using receiver operator characteristic (ROC) curve analysis in order to determine the area under the curve (AUC) as a measure of discriminative power. Patients were stratified based on the number of elevated MOCHA markers on admission. Sensitivity, specificity, positive predictive value (PPV), and negative predictive value (NPV) were evaluated at different MOCHA and D-dimer cutoffs. This was used to identify optimal thresholds for predicting thrombotic endpoints and to compare the performance of admission MOCHA with admission D-dimer alone. Statistical analyses and figures were generated using SPSS version 26 software.

## RESULTS

### Descriptive Data

A total of 297 confirmed COVID-19 patients were hospitalized during the study period and had MOCHA profile within 72 hours of admission. After exclusion of 18 patients on outpatient anticoagulation therapy and three pregnant patients, the analysis cohort included 276 patients.

The mean age of the cohort was 59 ± 6 years, 130 (47%) were female, and 160 (62%) were African American. Median BMI was 30 (IQR: 26-37) kg/m^2^ (Table 1). Common comorbidities included hypertension (59%), obesity (n=140, 51%) and diabetes (n=108, 39%). The median duration of hospitalization was 10 days (IQR: 6-19) including 159 patients (58%) admitted to the ICU during their hospitalization, 90 (33%) who were intubated. Amongst the cohort, 241 (87%) patients were discharged, 31 (11%) died and 4 (1%) remained hospitalized as of the censor date.

**Table 1.**
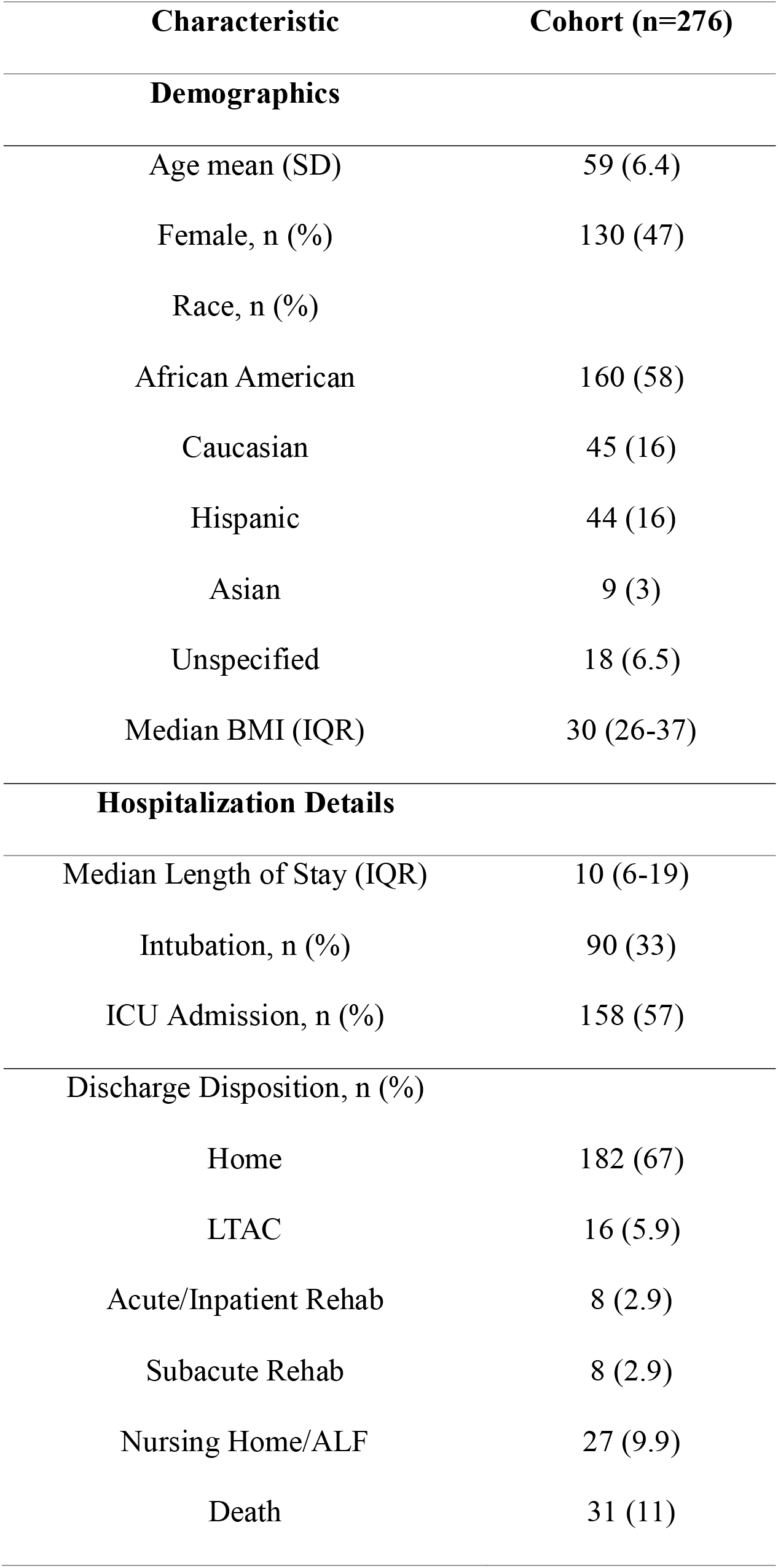
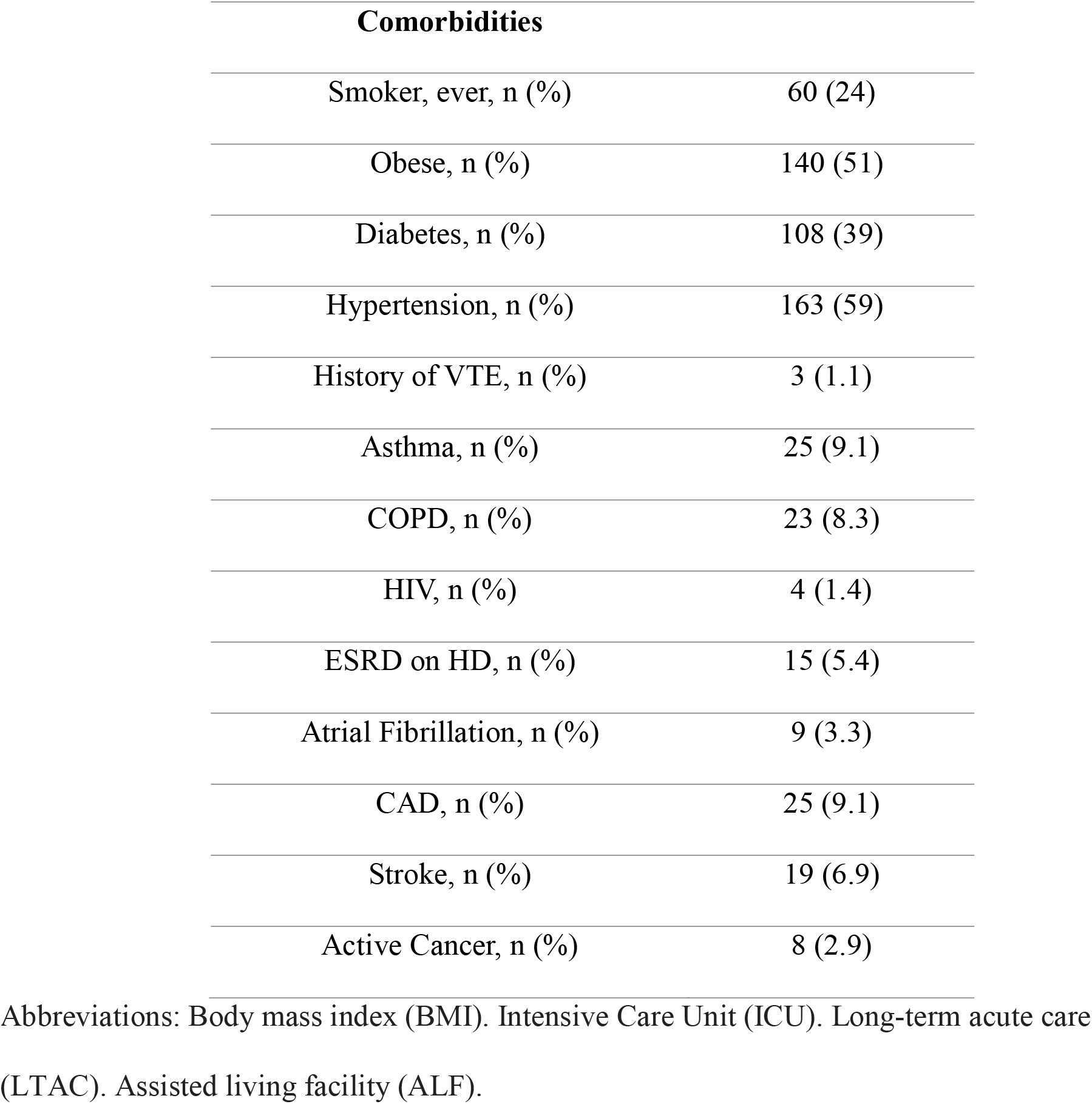
Patient characteristics and outcomes.

### Frequency of Vascular Events

Vascular events were diagnosed in 45 patients (16%) (Table 2). Median number of days to diagnosis of vascular event was 7 days (IQR: 2-15) from admission, with 32 (71%) of these events occurring within the first two weeks of hospitalization. DVT occurred in 24 (8.7%) patients, PE in 8 (2.9%), MI in 4 (1.5%), ischemic stroke in 5 (1.8%) and central or dialysis line thrombosis in 7 (2.5%) patients; three patients (1%) developed more than one of these complications.

**Table 2.**
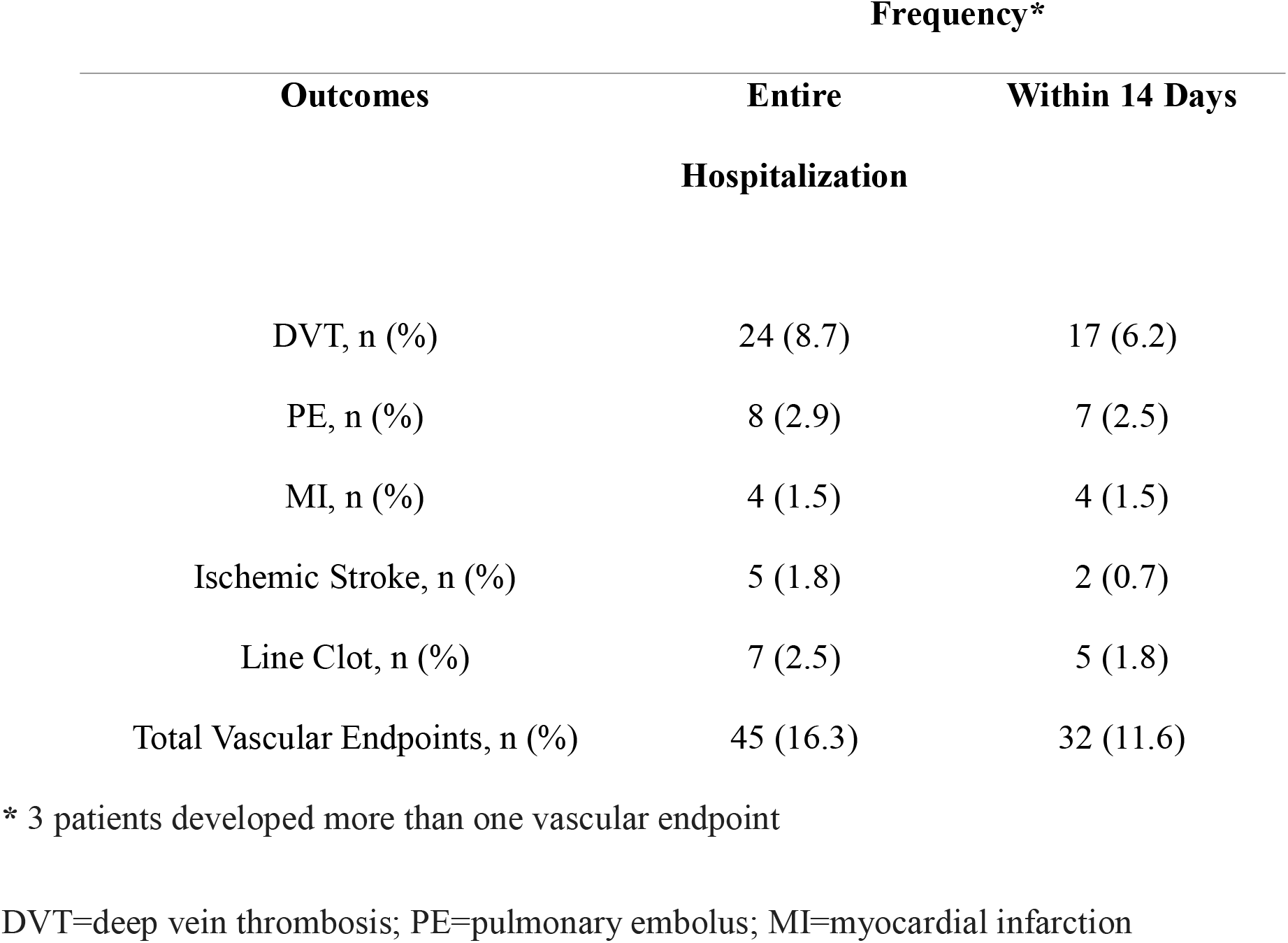
Frequency of Outcomes.

Overall, 7 (2.5%) patients had no MOCHA abnormalities on admission, 66 (24%) had one abnormality, 62 (23%) had two abnormalities, 69 (25%) had three abnormalities, and 72 (26.1%) had abnormalities in all four MOCHA; 217 patients (79%) had an abnormal D-dimer on admission, 51 (19%) had an admission D-dimer greater than 2000 ng/mL, and 40 (15%) had an admission D-dimer greater than 3000 ng/mL. There were 115 (42%) patients who had an elevated prothrombin fragment 1.2 level, 185 (67%) with an elevated thrombin-antithrombin complex level, and 167 (61%) with elevated fibrin monomer levels. The frequency of thrombotic events, ICU admission and intubation rates progressively increased with the number of MOCHA abnormalities (Figure 1).

**Figure 1.**
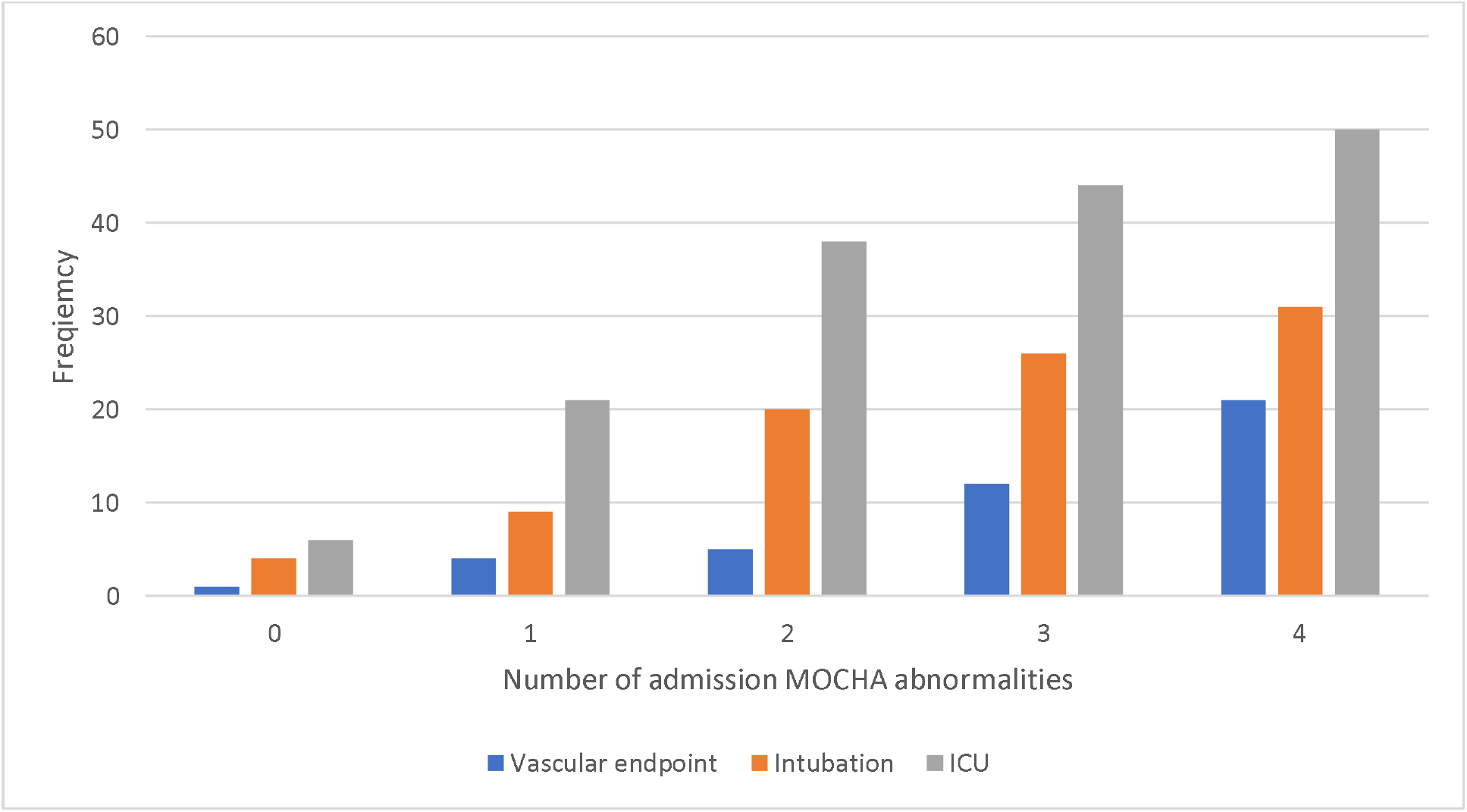
Frequency of MOCHA abnormalities in patients with vascular endpoints, intubation and ICU admission.

### Association of Admission MOCHA and D-dimer with Vascular Events

In univariable analysis admission MOCHA ≥ 2 abnormalities, D-dimer ≥ 2000 ng/mL, D-dimer ≥ 3000 ng/mL, BMI, and CAD were significantly associated with vascular events. However, in multivariable analysis only MOCHA ≥ 2 (OR 3.3, 95% CI 1.2-8.8; p=.02), BMI (OR 1.04, 95% CI 1.0-1.1; p=.03), D-dimer ≥ 2000 ng/mL (OR 3.1, 95% CI 1.5-6.6; p=.003), and D-dimer ≥ 3000 ng/mL (OR 3.6, 95% CI 1.6-7.9; p=.002) were significant. In multivariable analysis, factors associated with vascular events within 14 days of admission included male sex (OR 2.2, 95% CI 1.0-5.0; p=.049), MOCHA ≥ 2 abnormalities (OR 6.4, 95% CI 1.5-27.6; p=.013) and D-dimer ≥ 2000 ng/mL (OR 2.5, 95% CI 1.1-5.9; p=.03).

### Association of Admission MOCHA and D-Dimer with ICU Admission, Intubation and Mortality

In univariable analysis, admission MOCHA ≥ 2 abnormalities and a history of stroke were significantly associated with ICU admission. However, in multivariable analysis, only admission MOCHA ≥ 2 abnormalities was a significant predictor of ICU admission (OR 3.0, 95% CI 1.7-5.2; p=.0001).

With regards to the risk of respiratory deterioration and intubation, BMI, history of hypertension, and MOCHA ≥ 2 abnormalities were associated with intubation in univariable analysis. In multivariable analysis, MOCHA ≥ 2 abnormalities (OR 3.2, 95% CI 1.6-6.4; p=.001) and BMI (OR 1.04, 95% CI 1.0-1.1; p=.017) remained significant. D-dimer was not significantly associated with either ICU admission or intubation.

In univariable analysis, admission MOCHA ≥ 2 abnormalities, history of stroke, hypertension, diabetes and BMI were significantly associated with mortality. In multivariable analysis, only BMI remained a significant predictor of mortality (OR 1.04, 95% CI 1.01-1.08; p=0.02).

### Value of Admission MOCHA and D-dimer in Predicting Vascular Events, VTE, ICU Admission, and Intubation

Sensitivity, specificity, PPV, and NPV at different MOCHA and D-dimer cutoffs are shown in Table 3. A MOCHA profile with ≥ 2 abnormalities had a sensitivity of 89% (NPV 93%) for a vascular thrombotic event through total hospitalization, 94% (NPV 97%) for a vascular thrombotic event within two weeks of admission, and 96% (NPV 99%) for VTE within two weeks of admission. D-dimer level ≥ 2000 ng/mL on admission had a sensitivity of 33% (NPV 87%) for a vascular thrombotic event through total hospitalization, sensitivity of 31% (NPV 90%) for a vascular thrombotic event within two weeks of admission and a sensitivity of 43% (NPV 94%) for VTE within two weeks.

**Table 3.**
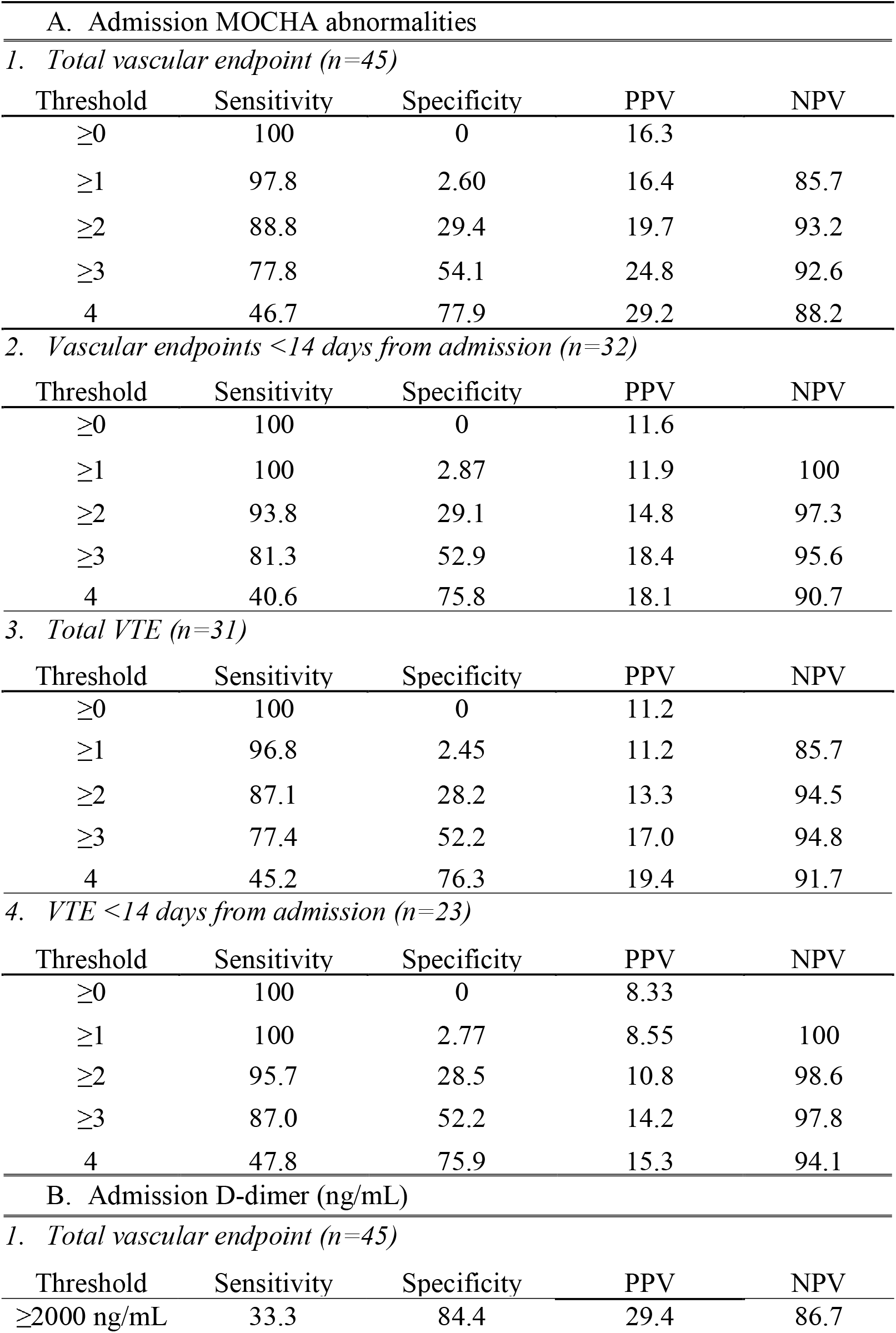
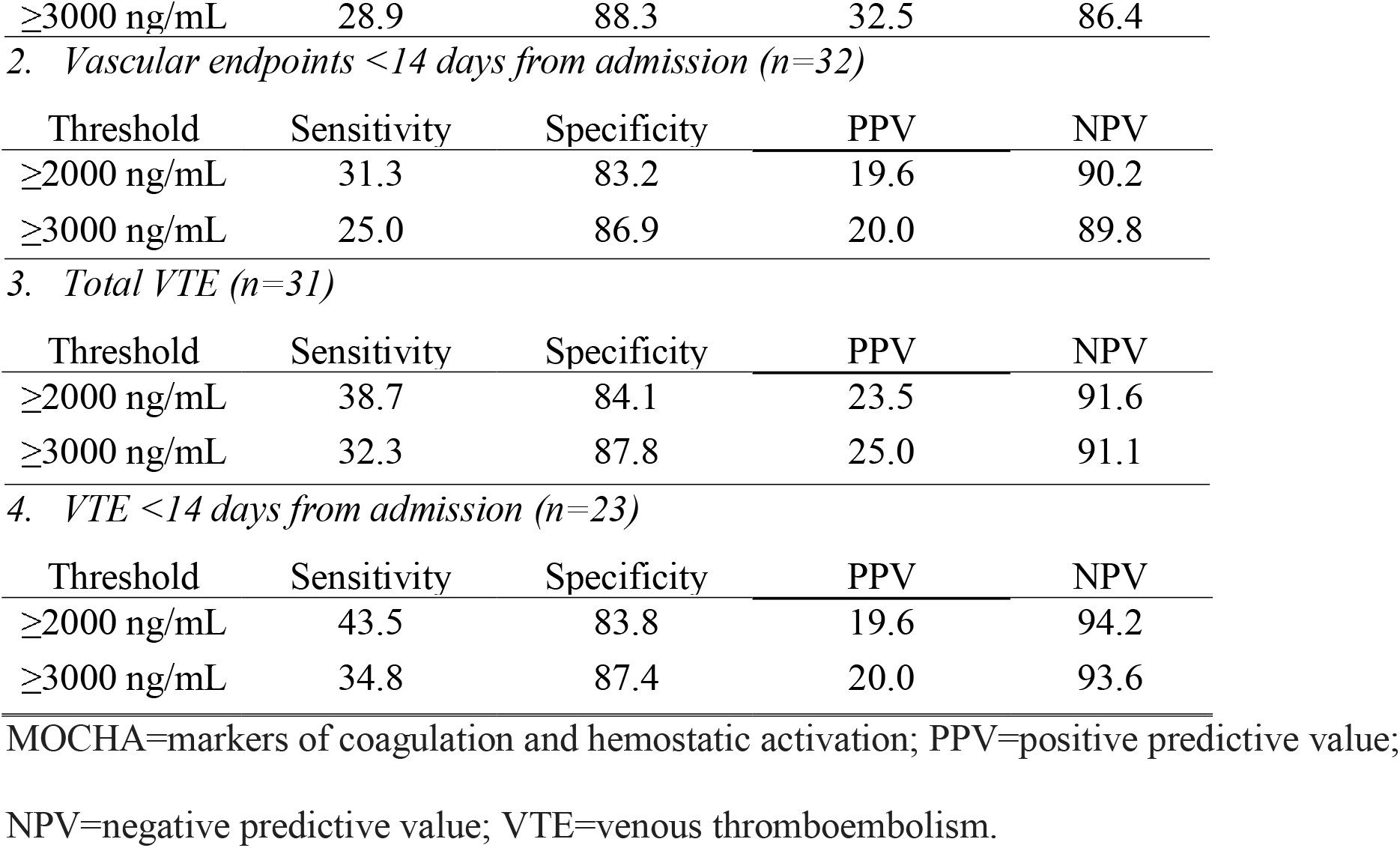
Sensitivity, specificity, positive predictive value and negative predictive value for (A) admission MOCHA and (B) D-dimer cutoffs with vascular endpoints.

| A. Admission MOCHA abnormalities |  |  |  |  |
| --- | --- | --- | --- | --- |
| 1. Total vascular endpoint (n=45) |  |  |  |  |
| Threshold | Sensitivity | Specificity | PPV | NPV |
| ≥0 | 100 | 0 | 16.3 |  |
| ≥1 | 97.8 | 2.60 | 16.4 | 85.7 |
| ≥2 | 88.8 | 29.4 | 19.7 | 93.2 |
| ≥3 | 77.8 | 54.1 | 24.8 | 92.6 |
| 4 | 46.7 | 77.9 | 29.2 | 88.2 |
| 2. Vascular endpoints <14 days from admission (n=32) |  |  |  |  |
| Threshold | Sensitivity | Specificity | PPV | NPV |
| ≥0 | 100 | 0 | 11.6 |  |
| ≥1 | 100 | 2.87 | 11.9 | 100 |
| ≥2 | 93.8 | 29.1 | 14.8 | 97.3 |
| ≥3 | 81.3 | 52.9 | 18.4 | 95.6 |
| 4 | 40.6 | 75.8 | 18.1 | 90.7 |
| 3. Total VTE (n=31) |  |  |  |  |
| Threshold | Sensitivity | Specificity | PPV | NPV |
| ≥0 | 100 | 0 | 11.2 |  |
| ≥1 | 96.8 | 2.45 | 11.2 | 85.7 |
| ≥2 | 87.1 | 28.2 | 13.3 | 94.5 |
| ≥3 | 77.4 | 52.2 | 17.0 | 94.8 |
| 4 | 45.2 | 76.3 | 19.4 | 91.7 |
| 4. VTE <14 days from admission (n=23) |  |  |  |  |
| Threshold | Sensitivity | Specificity | PPV | NPV |
| ≥0 | 100 | 0 | 8.33 |  |
| ≥1 | 100 | 2.77 | 8.55 | 100 |
| ≥2 | 95.7 | 28.5 | 10.8 | 98.6 |
| ≥3 | 87.0 | 52.2 | 14.2 | 97.8 |
| 4 | 47.8 | 75.9 | 15.3 | 94.1 |
| B. Admission D-dimer (ng/mL) |  |  |  |  |
| 1. Total vascular endpoint (n=45) |  |  |  |  |
| Threshold | Sensitivity | Specificity | PPV | NPV |
| ≥2000 ng/mL | 33.3 | 84.4 | 29.4 | 86.7 |
| $\geq 3000$ ng/mL | 28.9 | 88.3 | 32.5 | 86.4 |

2. *Vascular endpoints <14 days from admission (n=32)*
| Threshold | Sensitivity | Specificity | PPV | NPV |
| --- | --- | --- | --- | --- |
| $\geq 2000$ ng/mL | 31.3 | 83.2 | 19.6 | 90.2 |
| $\geq 3000$ ng/mL | 25.0 | 86.9 | 20.0 | 89.8 |

3. *Total VTE (n=31)*
| Threshold | Sensitivity | Specificity | PPV | NPV |
| --- | --- | --- | --- | --- |
| $\geq 2000$ ng/mL | 38.7 | 84.1 | 23.5 | 91.6 |
| $\geq 3000$ ng/mL | 32.3 | 87.8 | 25.0 | 91.1 |

4. *VTE <14 days from admission (n=23)*
| Threshold | Sensitivity | Specificity | PPV | NPV |
| --- | --- | --- | --- | --- |
| $\geq 2000$ ng/mL | 43.5 | 83.8 | 19.6 | 94.2 |
| $\geq 3000$ ng/mL | 34.8 | 87.4 | 20.0 | 93.6 |

- 1 MOCHA=markers of coagulation and hemostatic activation; PPV=positive predictive value; - 2 NPV=negative predictive value; VTE=venous thromboembolism.

ROC curve analysis showed that all four MOCHA parameters were independent predictors of any vascular endpoint (p<.01). However, the discriminative power for MOCHA was highest for VTE within the first two weeks of admission, during which time D-dimer and thrombin-antithrombin complex both had AUC 0.74, prothrombin fragment 1.2 had AUC 0.68 and fibrin monomer AUC 0.62.

## DISCUSSION

Our study provides systematic assessment of the admission MOCHA profile in hospitalized COVID-19 patients and its association with vascular thrombotic events. While the frequency of elevated D-dimer and prothrombin fragment 1.2 is similar to those reported in recent studies^7,12^, our study is the first to report the frequency of thrombin-antithrombin complex and fibrin monomer levels in hospitalized COVID-19 patients. While ≥ 2 abnormalities in the admission MOCHA profile and D-dimer cutoffs were significantly associated with vascular thrombotic events only the admission MOCHA profile was associated with subsequent ICU admission and intubation whereas D-dimer cutoffs of ≥ 2000 ng/mL and ≥ 3000 ng/mL were not.

We assessed these four markers of coagulation and hemostatic activation given their prior association with thrombotic events.^9-11^ D-dimer is a marker of fibrinolysis as a byproduct of fibrin degradation. Prothrombin fragment 1.2 is a marker of coagulation activation and released during conversion of prothrombin to thrombin. Thrombin-antithrombin complex is a marker of coagulation activation and a complex formed during thrombin formation. Fibrin monomer (soluble fibrin) is a marker of coagulation activation and a byproduct of fibrinogen conversion to fibrin.^14^

We found no association between admission MOCHA profile or admission D-dimer and mortality, contrary to early studies from China which suggested that D-dimer was a predictor of mortality in pooled analysis.^13^ There are several factors which may contribute to this difference: 1) our patients were all placed on prophylactic or therapeutic doses of anticoagulation therapy according to our local guidelines which may have influenced overall mortality rates which are lower in our cohort than other studies in the pooled analysis^8^; 2) the Chinese studies in this pooled analysis were all retrospective cohorts and excluded patients who were still hospitalized at the end of the study period contributing to a high risk of bias.^13^

Given current recommendations on the use of anticoagulation therapy in COVID-19 patients^15^, we also sought to identify whether the MOCHA profile could identify a COVID-19 patient subgroup that was at low risk for vascular thrombotic events. In our cohort, 26% of patients on admission had 0 or 1 MOCHA abnormality with NPV of 93% for vascular thrombotic events and NPV of 95% for VTE. These data suggest that this patient subgroup is at low risk for subsequent thrombotic complications.

This study had several limitations. As the data was collected from a single academic healthcare system, the generalizability of these findings needs to be studied in other settings. Our study focused on admission coagulation markers to aid in early risk stratification for COVID-19 patients however it is unknown whether serial measurements and changes in values across time may be better predictors. Lastly, our study did not assess whether the MOCHA profile could be used to guide anticoagulation therapy and how that may impact overall thrombotic events and clinical outcome.

In summary, the admission MOCHA profile of hospitalized COVID-19 patients is useful in identifying hospitalized patients who are at increased risk for subsequent arterial and venous thrombotic events and more effectively identifies patients requiring ICU admission and intubation than admission D-dimer levels alone. Further investigation is needed to determine the utility of the MOCHA profile to guide anticoagulation therapy.

## Data Availability

All data is available for approved requests made to the corresponding author.

